# Visual Outcome of Manual Small Incision Cataract Surgery in Patients with Diabetes Mellitus at the University of Gondar Tertiary Eye Care and Training Center

**DOI:** 10.1101/2025.08.21.25333501

**Authors:** Aschalew Mulugeta, Asamere Tsegaw, Wossen Mulugeta

## Abstract

**Background:** Manual Small Incision Cataract Surgery /MSICS/ is a widely used, cost-effective surgical technique for cataract removal and diabetic patients with cataract are high-risk groups who often experience poorer visual outcomes after cataract surgery due to various risk factors, including retinopathy progression. Hence, knowing the visual outcome and identifying factors affecting it, is critical for improving surgical success rates and patients’ quality of life.

**Objective:** To determine Visual Outcome of MSICS and Associated factors in patients with Diabetes and cataract at the University of Gondar Specialized Hospital Tertiary Eye Care and Training Center /UoGSH-TETC/, Gondar, North West Ethiopia.

**Methods:** A prospective study was conducted at the UoGSH-TETC, from January 2024 to December 2024. Data were collected on each follow-up visit, coded and analyzed using SPSS version 25. Descriptive statistics was used to summarize the data. Qualitative and quantitative data analysis was performed using Student’s *Paired t-*Test and Chi-square test respectively and *p-values* < 0.05 was considered as statistically significant. Outcome was measured by improvement in visual acuity (VA) from baseline preoperative acuity using electronic Snellen’s VA chart displayed on a computer screen, and WHO criteria for cataract surgery outcome was used as a reference for comparison.

**Results:** A total of 72 eyes of 69 patients completed post-operative follow up of 6 weeks and studied. The mean age at the presentation was 59.56 +/−12.12 years. Mean Best Corrected Visual Acuity /BCVA/ of 0.38+/−0.21 logMAR achieved at 6^th^ post-operative week from pre-operative mean VA of 1.53+/−0.47, p< 0.001. At 6^th^ post-operative week, 75% and 84.7% eyes had uncorrected visual acuity and BCVA of 6/18 or better, respectively and 62.5% had ≥6/12. At the final follow-up examination 11/15.3%/ eyes had various degrees of progression in their diabetic retinopathy from the baseline level.

**Conclusion and Recommendation:** This study showed that MSICS in diabetes patients with cataract resulted in an overall improvement in visual acuity and about 85% of operated patients archived good visual outcome after refraction. Although this finding is slightly below the WHO recommendations, it is still better than previous similar cataract surgical outcomes studies done at the same place and other sites in the country.

## Introduction

WHO defines Cataract as clouding of the crystalline lens of the eye. It is the leading cause of preventable blindness worldwide.1,2 Diabetes mellitus (DM) is defined as a group of metabolic diseases characterized by hyperglycemia resulting from defects in insulin secretion, insulin action, or both.3 Its prevalence is increasing on a daily basis with the International Diabetes Federation estimating that there will be 439 million DM patients by 2030. 4,5,6 The development of cataract is the second most common ocular complication of DM. 6 The main risk factors for cataract development in DM patients are longer duration of diabetes and poor metabolic control. 7 Due to the increasing prevalence of DM, the incidence of diabetic cataracts has also risen. 7

Cataract surgery is the removal of an opacified lens and is usually replaced with an artificial intraocular lens (IOL), and it is the first and last available management of cataracts. A better understanding of various factors responsible for favorable outcomes of cataract surgery in diabetic patients may guide us in better overall management of these patients and optimizing the results.7,8,9

Cataract surgery in diabetics is indicated for visual improvement, allowing diagnosis/ assessment and treatment of DR and maculopathy. Small Incision Cataract Surgery (SICS) is a widely used cost-effective surgical technique for cataract extraction, particularly in regions with limited access to more advanced surgical methods. However, cataract surgery in a diabetic patient is associated with several difficulties and carries a higher risk of both intra-and post-operative complications compared to non-diabetics. 6, 11,12,13,14

Recent studies on cataract surgery in diabetics tend to report a lower incidence of complications and better visual outcomes due to better preoperative management of retinopathy, evolutions in operative techniques, and better glycemic and hypertensive control.8,15 World Health Organization (WHO) recommends that after surgery at least 80% of the operated eyes should have a presenting visual acuity of 6/6–6/18, which is referred to as a good visual outcome whereas poor visual outcome cases (less than 6/60) should have less than 5%. After the best correction, at least 90% of the eyes should achieve good visual outcome.

Post-operative visual acuity is the most popular way to assess the quality of cataract surgery. 8,9,15 Very few large-scale studies have evaluated the visual outcomes of people with diabetes after cataract surgery.

The aim of this study, then, was to determine the visual outcome following MSICS and factors affecting the outcome in diabetic patients with diabetes at the University of Gondar comprehensive specialized hospital Tertiary eye care and training center.

## METHODS AND MATERIALS

### Study Design and Period

A hospital-based prospective follow-up study was conducted in all diabetic cases with cataract who meet the inclusion criteria, at UoG Comprehensive Specialized Hospital Tertiary Eye Care and Training Center, from January 2024 to December 2024.

### Study Area

The study was conducted at the University of Gondar (UoG) Tertiary Eye Care and Training Center, a major eye care and training center in Ethiopia. It is an ophthalmic referral center for an estimated 14 million people living in North-West Ethiopia. The center provides eye care services both at the base eye center and rural outreach sites. The base eye center has 8 outpatient clinics, facilities for in-patient care with 30 beds and four rooms with beds for eye surgery (operation theatres). At the time of this study, the center had 8 ophthalmologists, 6 of them sub-specialists. There were also 26 Ophthalmology residents, over 30 optometrists and ophthalmic nurses, and 29 general clinical nurses actively working in the outpatient clinics and operation theatres of the Tertiary Eye Care and Training Center. On average, up to 50,000 patients are seen as an outpatient and over 5000 major eye surgeries done at the base eye center annually.

### Study Population

All diabetic patients with cataract who underwent MSICS and fulfilled the inclusion criteria

### Inclusion criteria

All patients having known DM with complete or partial opacification of the lens which impaired vision and those who undergone cataract surgery during the study period.

### Exclusion criteria

Patients with other causes of reduction of vision, such as glaucoma, AMD, and corneal opacities etc.

Patients having poorly controlled glycemic status (FBS level of >200 mg/dl or HbA1c of >8.5%).

Patients with any secondary cataract other than DM (e.g., traumatic Cataract, uveitic cataract etc…).

Patients who lost from follow-up after surgery before completing the required 6 weeks post operative follow up period.

### Sample Size and Sampling Procedure

Consecutive sampling technique was used to include diabetic patients who undergone cataract surgery during the study period.

### Surgical Procedure of MSICS

After IOL power calculation (biometry), informed written consent was taken from the patient and the eye was prepared for surgery including eye lash trimming, dilating the pupil with tropicamide 1% (with or without phenylephrine), and labeling the eye to be operated. Local anesthesia injection was given with lidocaine 2% 5ml (with or without adrenaline) to achieve anesthesia and akinesia of the operated eye and periocular structures. 5% povidone iodine solution was instilled to the conjunctival fornix in the operating theater prior to the surgery. The periocular area was cleaned with 10% povidone iodine, the eye draped and speculum inserted.

After conjunctival peritomy at the temporal limbus, the underlying sclera was exposed and cauterized. A crescent blade was used to create a frown-shaped partial-thickness scleral incision approximately 2 mm posterior to the limbus, extending laterally for approximately 6-7 mm. This incision was beveled to facilitate the creation of a self-sealing tunnel. Following this, dissection was carefully performed within the scleral tissue to create a scleral tunnel of 3-4 mm length, using the same crescent blade. A keratome was then used to enter the anterior chamber (AC). After AC entry, visco-elastics was injected to maintain the AC depth and protect corneal endothelium.

After can-opener capsulorhexis, hydro-dissection was done and nucleus prolapsed into the AC and removed through the scleral tunnel using a lens loop. Any residual cortical material was washed using a Simcoe cannula and PMMA intraocular lens (IOL) implanted into the capsular bag. The AC was then reformed with a balanced salt solution. No suture was required as the tunnel was self sealing. At the end of the procedure, subconjunctival injection of Dexamethasone mixed with Gentamycin was given. Subconjunctival Triamcinolone Acetonide injection was also routinely given for all patients as additional anti-inflammatory agent.

Our routine post operative eye medications were Dexamethasone 0.1% eye drop applied 2hourly for the first week and four times a day for the next 3 weeks and ciprofloxacin 0.3% eye drop four times a day for the first postoperative week.

Operated patients were examined on the first day, first week and on the 6^th^ week after surgery. Spectral domain-optical coherence tomography, SD-OCT (SD; Cirrus HD, Carl Zeiss Meditec, USA) scan of the retina was done for all patients on the 1^st^ and 6^th^ post operative weeks. Refraction was done after the 6^th^ postoperative week.

### Operational Definition

**Diabetes Mellitus**: diagnosis of DM is made when one of the three **criteria** was met. 3

1. Symptoms of diabetes plus casual plasma glucose concentration ≥ 200 mg/dl. Casual is defined as any time of day without regard to time since the last meal. The classic symptoms of diabetes include polyuria, polydipsia, fatigue, and unexplained weight loss.
2. Fasting plasma glucose ≥ 126 mg/dl. Fasting is defined as no caloric intake for at least 8 hours.
3. 2 hours plasma glucose ≥ 200 mg/dl during an oral glucose tolerance test. 3

**Type-I Diabetes Mellitus:** is characterized by β-cell destruction due to an autoimmune process leading to absolute insulin deficiency and diagnosed in people age less than 30 years old. 3

**Type 2 Diabetes Mellitus:** is characterized by insulin resistance in peripheral tissues and an insulin secretory defect of the β cells and its diagnosed in people older than 30 years. 3

**Non-Proliferative Diabetic Retinopathy (NPDR):** Describes intraretinal microvascular changes, which include basement membrane thickening, pericyte loss, microaneurysm formation, venous caliber abnormalities, retinal capillary closure, and intraretinal microvascular abnormalities (IRMAs). 16

**No diabetic retinopathy:** No retinal lesions characteristic of diabetic retinopathy seen
**Mild NPDR**: The presence of only retinal microaneurysms
**Moderate NPDR**: More than just microaneurysms but less than Severe NPDR

**Severe NPDR**: the presence of any one of the following features (4:2:1 rule) and no signs of proliferative retinopathy:

1. Severe (More than 20) intraretinal hemorrhage in each of 4 quadrants
2. Definite venous beading in 2 or more quadrants
3. Prominent intraretinal microvascular abnormalities (IRMA) in at least 1 quadrant **Very severe NPDR**: The presence of two or more of the above-mentioned features described in severe NPDR

**Proliferative Diabetic retinopathy (PDR)**: One or more of the following: Neovascularization over the disc (NVD) or on the retina (NVE), Vitreous/ pre-retinal hemorrhage.

**PDR with high-risk** characteristics is defined as having any of the following findings:

1. Any neovascularization over the disc (NVD) with vitreous or pre-retinal hemorrhage
2. Extent of NVD greater than or equal to one-fourth of disc area, with or without vitreous or pre-retinal hemorrhage
3. Extent of NVE greater than or equal to one-half disc area, with vitreous or pre-retinal hemorrhage

**Cataract** is defined as a painless disease of the eye in which a normally clear crystalline lens becomes cloudy or opaque, causing vision loss or reduction that cannot be corrected with glasses, contact lenses, or corneal refractive surgery. 17 Opacities are separated into four major groups: nuclear cataract (NC), cortical (CC), posterior subcapsular (PSC), and mixed (including anterior subcapsular). 18

**Types of complications after cataract surgery:** 19

- **No complication**
- **Posterior capsule rupture with/ without vitreous loss:** ruptured posterior lens capsule with/ without any vitreous loss
- **Intraocular lens (IOL) Capture:** entrapment of part of the IOL optic with the iris
- **Retained lens matter**: any clearly visible remains of the lens, apart from the anterior capsule, present in the eye after cataract surgery.
- **Striate keratopathy**: post operative corneal edema due to endothelial damage
- **Post operative Endophthalmitis**: intra-ocular infection, proven or suspected, following intraocular surgery

**Evaluation of Results** 19

**Presenting vision:** Visual acuity in the examined eye with the available correction, if any
**‘Best’ vision**: Visual acuity in the examined eye with the best possible correction or pinhole.

**WHO classification of acceptable visual acuity for cataract surgery outcome at 6th week:** 19

**Good Outcome:** When BCVA after cataract surgery is 6/18 or better (or VA≥20/60; VA≤0.5 (LogMar)).
**Borderline Outcome:** When BCVA after cataract surgery is <6/18–6/60.
**Poor Outcome:** When BCVA after cataract surgery is worse than 6/60 (<6/60 –LP) (or VA<20/200; VA>1.0 (LogMar).

**WHO recommendation of visual outcome after cataract surgery.** 19

- UCVA of good outcome in > 80% of the operated eyes at 6 weeks, borderline in < 15% and poor outcomes in only 5%, and
- BCVA of good results in > 90% of eyes, < 5% with borderline and < 5% with poor results, up to 6 weeks of post-surgery.

**Diabetic Retinopathy progression** is defined as:20,21

1. The development of any retinopathy where none previously existed, or worsening of the DR whether or not cystoid macular edema (CME) is present,
2. The development of proliferative retinopathy from NPDR or the recurrence of active PDR, and
3. The development of or progression of macular edema.

**Post operative Cystoid Macular edema (CME)** is commonly defined as a VA loss of 5 or more letters between postoperative day 7, when clinical CME is rare, and week 6 when most CMEs develop, in combination with macular changes on OCT, i.e., loss of foveal depression, and cystoid spaces within macula.22

### Data collection procedure

A structured questionnaire was used to collect sociodemographic data which included age, sex, residence, marital status, occupation etc and details of systemic clinical data which included duration and type of diabetes, type of treatment for the diabetes, recent fasting blood sugar level, any systemic comorbidity etc…. Preoperative ocular clinical data taken included the eye operated, preoperative visual acuity, preoperative status of the retina or any grade of diabetic retinopathy detected or not, type of cataract, power of calculated IOL implanted etc. Intraoperative and post operative ocular clinical data taken included incision site, intraoperative and post operative complication, post operative visual acuity at the first day, first week and 6^th^ week after surgery, best corrected visual acuity and post operative refractive error after refraction at the 6^th^ week, post operative status of the retina or level of diabetic retinopathy using clinical examination and OCT etc.

Preoperative and post operative VA was taken using electronic Snellen visual acuity chart displayed on a computer screen. Patients undergone detailed dilated slit-lamp examination both preoperative and post operative by ophthalmologists and findings recorded on each follow up day. Ocular Biometry or IOL power calculations and Refraction at the 6^th^ post operative week were done by senior optometrists. Both presenting and best corrected preoperative and post operative VAs was taken at an electronic Snellen’s VA chart desplayed on a computer screen by ophthalmologists.

Optical coherence tomography (OCT) was done on the 1^st^ and 5^th^ postoperative weeks for development or progression of any pathology after surgery. The OCT scans and dilated retinal examinations were performed by the Retina subspecialist, and finding were documented. All cataract surgeries were performed by a single ophthalmologist who is also a vitreo-retina subpsecialist.

### Data Processing and Analysis

The collected data was coded and entered into Epi-Data 4.6 and exported to Statistical Package for Social Sciences (SPSS) version 25 for further analysis. Descriptive analysis were made and a two-sided *p* value < 0.05 was considered to be statistically significant. The *t-test* for paired samples was performed to compare mean VA before and after surgery and results were described in terms of percentages, numbers, and means, then displayed on tables, graphs, and pie charts.

### Ethical Considerations

Ethical clearance was obtained from UoG, School of Medicine Ethical Review Committee (**letter of permission from ethical board needed or its refrence number cited here)**, and permission to undergo this study was obtained from the Department of Ophthalmology. Informed written consent was obtained from each study patient after a full explanation about the study. The patients data was used only for this study, confidentiality respected and was conducted as per the Helsinki declarations (reference).

## RESULTS

### Socio-demographic and Clinical Characteristics

Out of 85 diabetic patients operated for cataract using MSICS techniques during the study period, 69 of them (72 eyes) or 81% completed the required 6 weeks follow up period and studied. The mean age of study patients at presentation was 59.56 ± 12.12 (Range 24-84) years and 50.7% (35/69) were male. (Table-I)

**Table-I.**
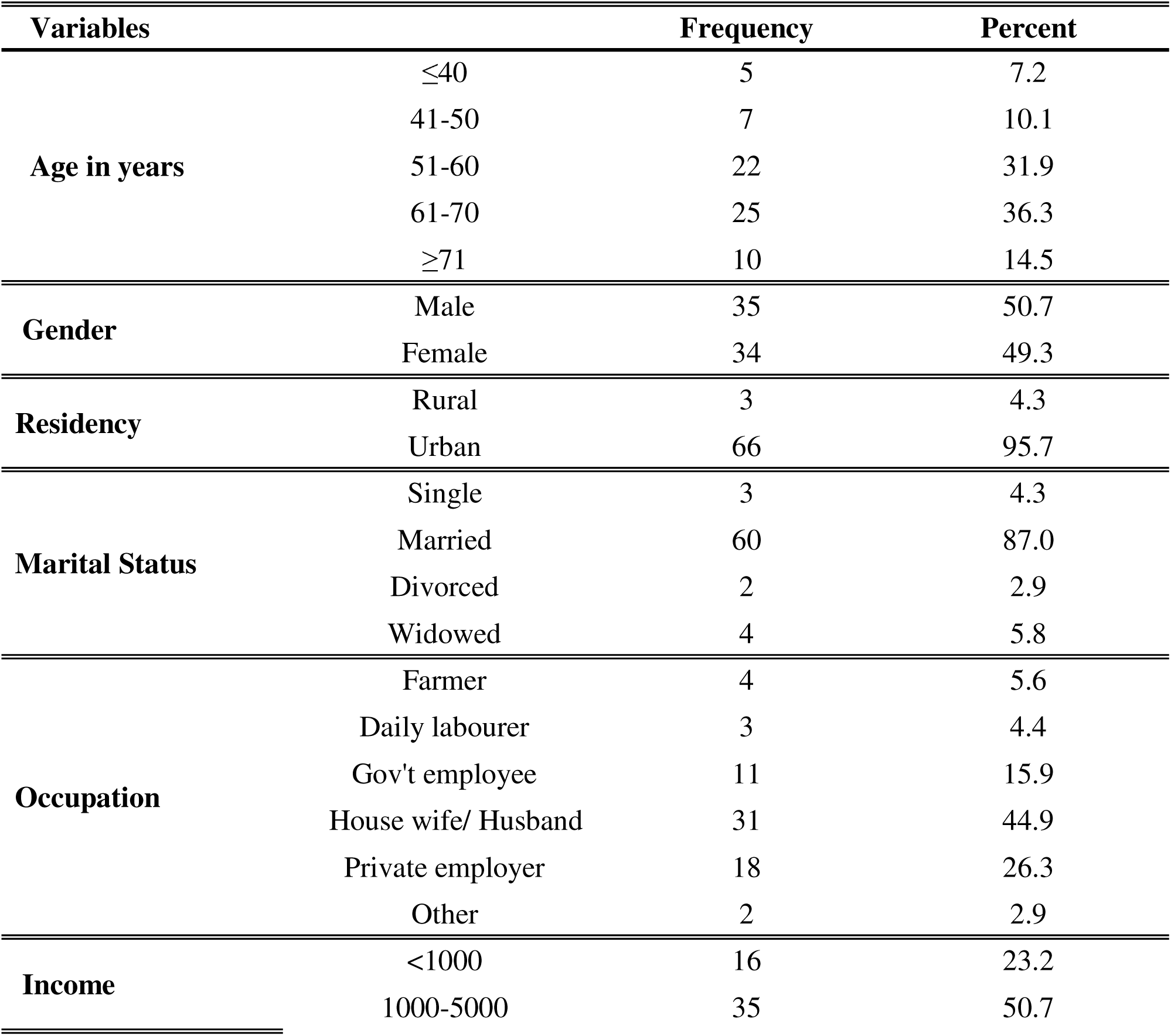
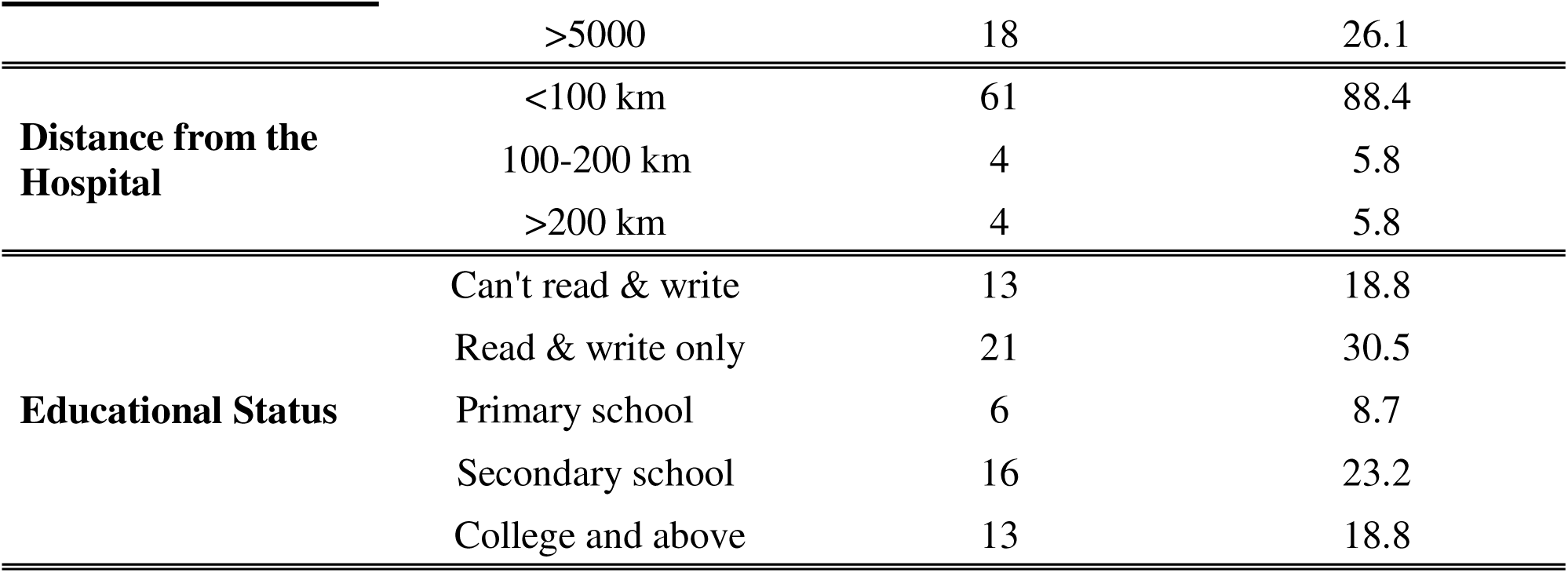
The socio-demographic characteristics of diabetic patients with cataract who underwent MSICS at the University of Gondar (UoG) Comprehensive Specialized Hospital: January -December, 2024.

**Table-II.** The clinical characteristics of diabetic patients who undergone MSICS at the UoG Comprehensive Specialized Hospital, January-December 2024.

| **Variables** |  | **Frequency** | **Percent** |
| --- | --- | --- | --- |
| **Type of Diabetes** | Type 1 | 7 | 10 |
|  | Type 2 | 62 | 90 |
| **Duration of Diabetes in years** | ≤5 | 17 | 24.7 |
|  | 6-10 | 26 | 37.7 |
|  | 11-19 | 15 | 21.7 |
|  | ≥20 | 11 | 15.9 |
| **Type of Treatment** | Diet & life style Modification | 3 | 4.3 |
|  | Oral medications | 32 | 46.4 |
|  | Insulin | 13 | 18.8 |
|  | Insulin & Oral medications | 21 | 30.5 |
| **FBS \* level in mg/ dl** | 65-100 | 15 | 20.8 |
|  | 101-140 | 37 | 51.4 |
|  | 141-179 | 20 | 27.8 |
| **Type of Cataract** | Involves all layers | 45 | 62.5 |
|  | PSC | 22 | 30.5 |
|  | Cortical | 4 | 5.6 |
|  | Nuclear | 1 | 1.4 |
| **Systemic Comorbidity** | Hypertension | 43 | 62.3 |
|  | Kidney Disease | 2 | 2.9 |
|  | Cardiac Disease | 3 | 4.3 |
|  | Stroke | 3 | 4.3 |
|  | RVI * | 2 | 2.9 |
| **BMI \* in Kg/ m<sup>2</sup>** | <18.5 | 2 | 2.9 |
|  | 18.5-24.9 | 39 | 56.5 |
|  | 25-29.9 | 27 | 39.1 |
|  | >30 | 1 | 1.5 |

\* FBS (fasting blood sugar), BMI (body mass index), PSC (posterior subcapsular cataract), RVI (retro-viral infection).

A majority, 62 (90.0%), of study patients had type II diabetes mellitus and the mean duration of diabetes at presentations was 10.81± 6.47 (range 4 months to 26) years. Oral hypoglycemic agents was the method of treatment in 32 (46.4%) patients, followed by combined insulin and oral medication in 21 (30.5%), then insulin by 13 (18.8%) patients. The commonest systemic comorbidity found was hypertension, seen in 43 (62.3%) patients. Over 83.3% of diabetes with retinopathy changes had hypertension. Additionally, 27 (39.1%) patients were found to be over-weight (BMI: 25-29.9 kg/m2).

The mean FBS level just before surgery was 125.5 ± 26.84 mg/dl (Range: 65-179). All of MSICS were done through temporal approach by a single ophthalmologist who is also the second author of this study.(Table 2)

**Table 1.** Distribution of first postoperative day visual acuity (VA) of diabetic patients who undergone MSICS at UoG Comprehensive Specialized Hospital Tertiary Eye care and training center, January to December 2024.

| Uncorrected VA on the first postoperative Day |  |  |
| --- | --- | --- |
| Visual Acuity | No. of eyes | Percentage |
| <6/60 | 33 | 45.8 |
| 6/24-6/60 | 20 | 27.8 |
| 6/15-6/18 | 10 | 13.9 |
| 6/6-6/12 | 9 | 12.5 |

**Table 2.** Distribution of Uncorrected visual Acuity of diabetic patients with cataract at the 6th week post MSICS at UoG Comprehensive Specialized Hospital Tertiary Eye care and training center, January to December 2024.

| **Uncorrected VA at the 6<sup>th</sup> postoperative week** |  |  |  |
| --- | --- | --- | --- |
|  | **Visual Acuity** | **No. of eyes** | **Percentage** |
| **Poor Outcome** | <6/60 | 1 | 1.4 |
| **Borderline** | <6/18-6/60 | 17 | 23.6 |
| **Good Outcome** | 6/6-6/18 | 54 | 75 |

The mean diopteric (D) difference in the implanted and calculated intra-ocular lens (IOL) power was +0.21D±0.29 (Range: +0.00 to +1.50 D). There was no IOL power difference between calculated and implanted in 46 (63.9%) eyes, but about one-third of [24 (33.3%)] eyes had +0.50 D difference. Preoperative ocular comorbidity other than DR was detected in 6 (8.3%) eyes including Myopic fundus, seen in 2 (2.8%) eyes, Pseudoexfoliation in 2 (2.8%), and Asteroid hyalosis and Posterior vitreous detachment (PVD) each seen in 1 (1.4%) eye.

Refraction was done in 41 (56.9%) eyes at 6^th^ postoperative week and refractive error was detected in 25 (61%) of them. Myopia was seen in 16 (39%) eyes; most of which [13 (81.3%)] were low-myopia, and Hyperopia was found in 3 (7.3%) eyes. Astigmatism was detected in 6 (14.6%) eyes, with 4 (66.7%) simple-myopic astigmatism (SMA) and 2 (33.3%) compound myopic astigmatism (CMA).

### Visual Outcomes of the Surgery

Both preoperative and postoperative VA was recorded using electronic Snellen’s VA chart and the values were converted to log MAR units for statistical analysis. The mean preoperative VA in log MAR was 1.53±0.47 (Range: 0.8-2.7), and after sixth postoperative weeks it changed to UCVA of 0.45±0.25 and BCVA of 0.38±0.21, p < 0.001 showing final visual outcome has improved significantly after MSICS (Figure 1). Uncorrected visual acuity on the first postoperative day (Table 3) was poor (<6/60) in 33 (45.8%) eyes, mostly due to corneal edema. But UCVA on the sixth week of follow up was ≥6/18 in 54 (75%) eyes (Table 4) and the majority of eyes, 61 (84.7%), had BCVA of 6/18 or better (Table 5) and 45 (62.5%) had VA of ≥6/12 (Figure 2).

**Figure 1.**
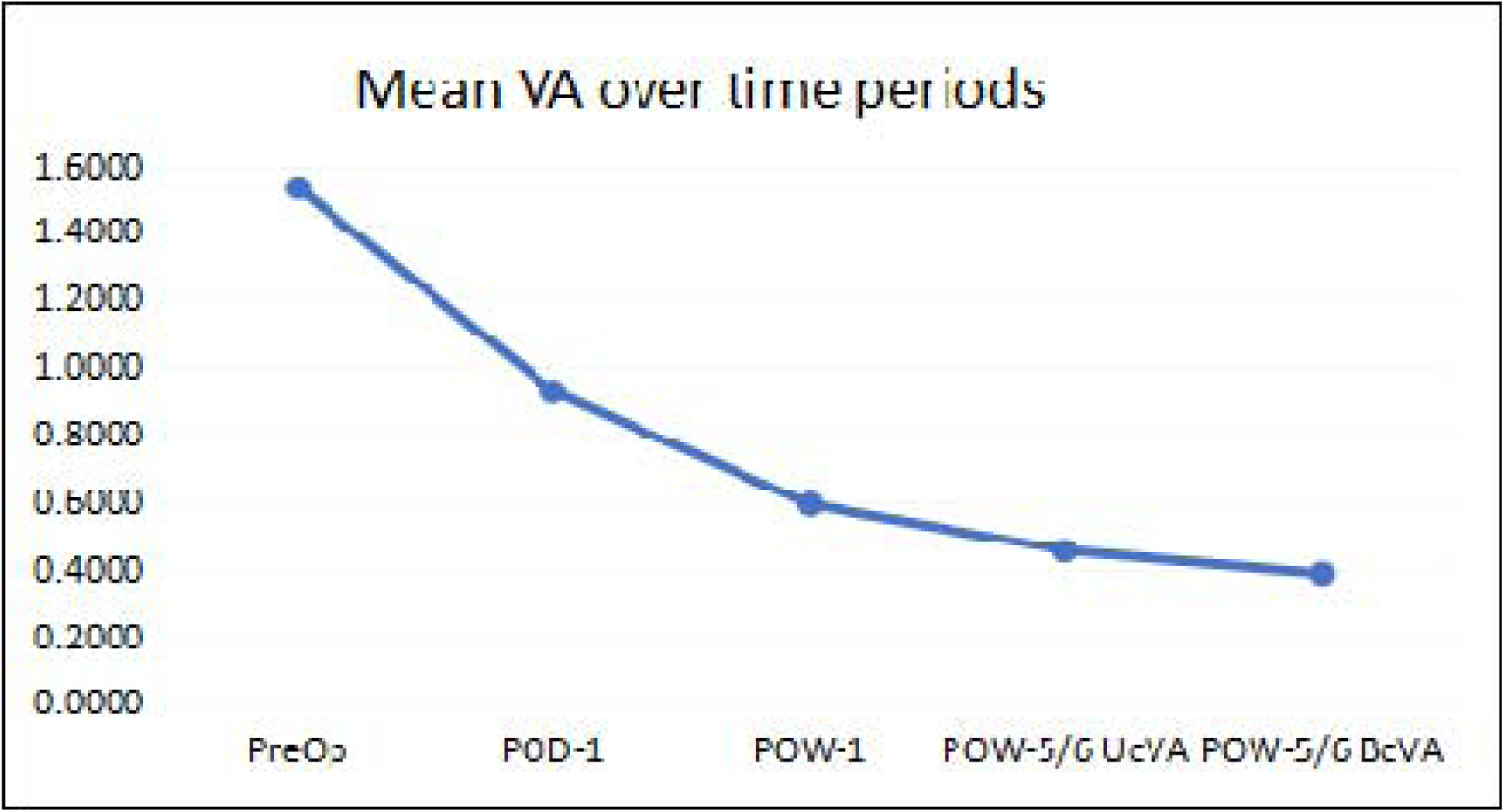
Graph showing pattern of change in the mean VA over the study period in logMAR unit at the UoG Comprehensive Specialized Hospital, tertiary eye care and training center, northwest Ethiopia, January 2024 to December 2024. *PreOp (preoperative VA), POD-1 (1^st^ postoperative day VA), POW-1 (1^st^ postoperative week VA), POW-5/6 UcVA (uncorrected VA on postoperative week-5 or 6), POW-5/6 BcVA (best-corrected VA on postoperative week-5 or 6), UoG (University of Gondar)*.

**Figure 2.**
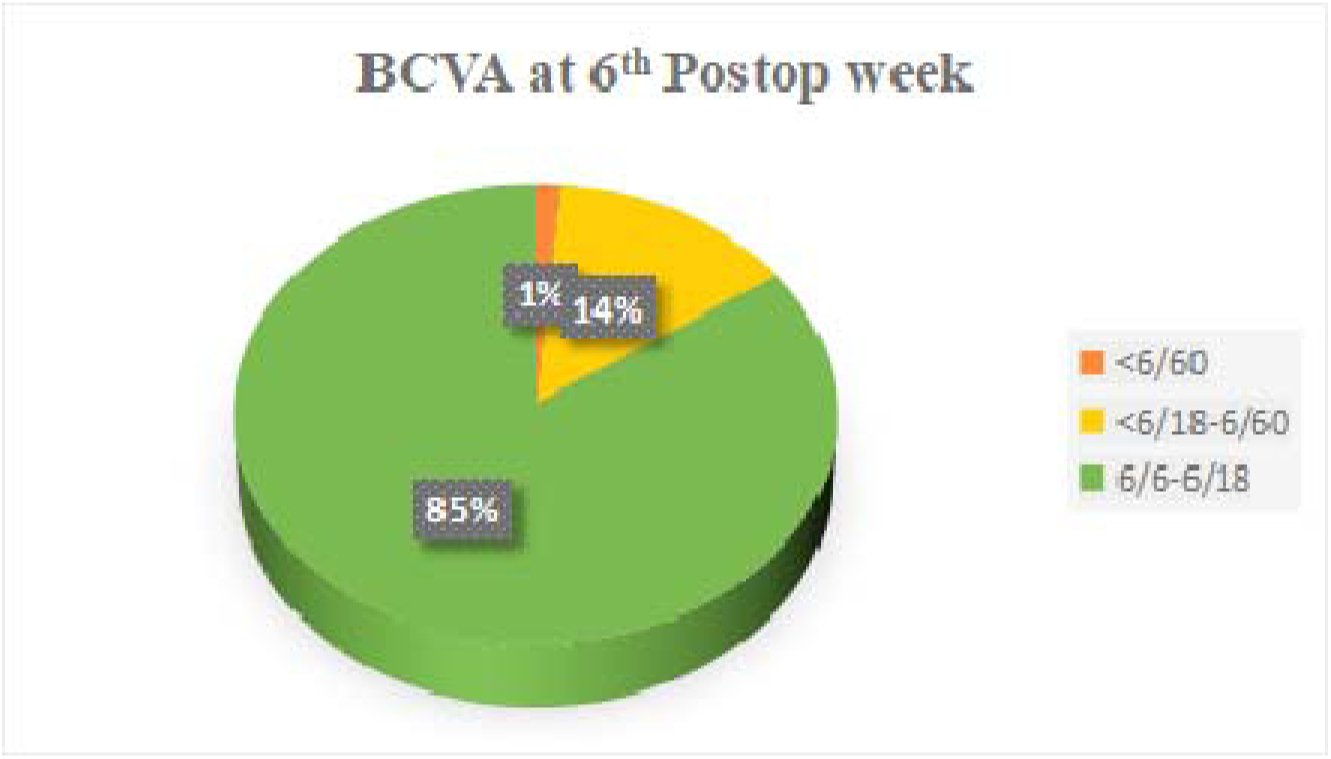
Graph showing the pattern of final visual acuity (6^th^ postoperative week) of diabetic patients with cataract at the Snellens acuity chart after SICS was done at the UoG Specialized Hospital Tertiary Eye care and training center, January to December 2024.

**Table 3.** Distribution of final visual outcome of diabetic patients with cataract who undergone MSICS at UoG Comprehensive Specialized Hospital Tertiary Eye care and training center,January to December 2024.

| **Best corrected VA at the 6<sup>th</sup> postoperative week** |  |  |  |
| --- | --- | --- | --- |
|  | **Visual Acuity** | **No. of eyes** | **Percentage** |
| **Poor Outcome** | <6/60 | 1 | 1.4 |
|  | <6/18-6/60 | 10 | 13.9 |
| **Borderline** | 6/6-6/18 | 61 | 84.7 |
| **Good Outcome** |  |  |  |

**Table 4.** Frequency distribution of intraoperative and postoperative Complications after MSICS in diabetic patients at the UoG Comprehensive Specialized Hospital, January to December 2024.

| **Complications** |  | **Frequency** | **Percent** |
| --- | --- | --- | --- |
| **Intraoperative** | Posterior capsular tear | 6 | 8.3 |
|  | No Complication | 66 | 91.7 |
| **Postoperative** | Corneal Edema/ Straight keratopathy | 18 | 25.0 |
|  | Retained lens material | 6 | 8.3 |
|  | Pupillary membrane | 3 | 4.2 |
|  | Cystoid Macular edema | 8 | 15.4 |
|  | Raised intraocular pressure | 2 | 2.8 |
|  | Anterior chamber inflammation | 2 | 2.8 |
|  | Posterior capsular opacity | 3 | 4.2 |
|  | Intraocular Lens Capture | 1 | 1.4 |

**Table 5.**
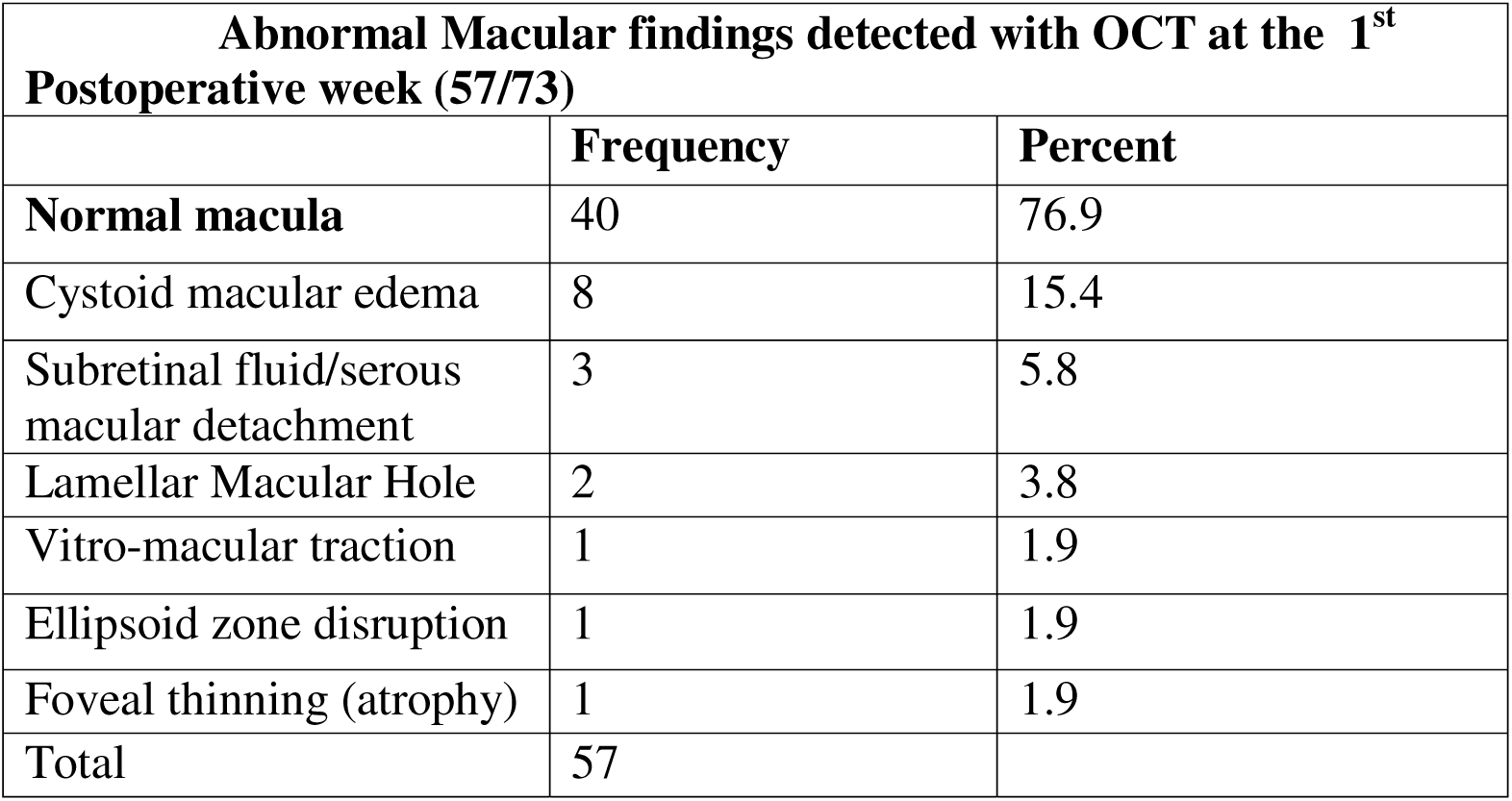
Patterns of 1^st^ week post operative abnormal macular OCT findings detected in diabetic patients with cataract after MSICS at the UoG Comprehensive Specialized Hospital, January to December, 2024.

| **Abnormal Macular findings detected with OCT at the 1<sup>st</sup> Postoperative week (57/73)** |  |  |
| --- | --- | --- |
|  | **Frequency** | **Percent** |
| **Normal macula** | 40 | 76.9 |
| Cystoid macular edema | 8 | 15.4 |
| Subretinal fluid/serous macular detachment | 3 | 5.8 |
| Lamellar Macular Hole | 2 | 3.8 |
| Vitro-macular traction | 1 | 1.9 |
| Ellipsoid zone disruption | 1 | 1.9 |
| Foveal thinning (atrophy) | 1 | 1.9 |
| Total | 57 |  |

Over all, the final BCVA in Snellen’s chart improved in one or more lines in 70 (97.2%) eyes, remained the same in 1(1.4%) and was worse in 1(1.4%) eye. There was only one patient who had poor visual outcome with final VA of light perception (LP) due to high-risk proliferative diabetic retinopathy complicated with vitreous hemorrhage and neovascular glaucoma.

A one-way ANOVA test (F-test) was used to compare the mean final BCVA based on the type of treatment for their diabetes and was found to be statistically significant (p = 0.023). Then, Tukey’s *Post-hoc* tests were used to determine the significant pair(s) and found that there was statistically significant difference in final visual outcome between patients who were on oral medications and those on Insulin (p= 0.024). There was no significant difference in final visual outcome based on the duration or type of diabetes, FBS level, BMI level, type of cataract, and demographic variables, p>0.05. (Figure 3)

**Figure 3.**
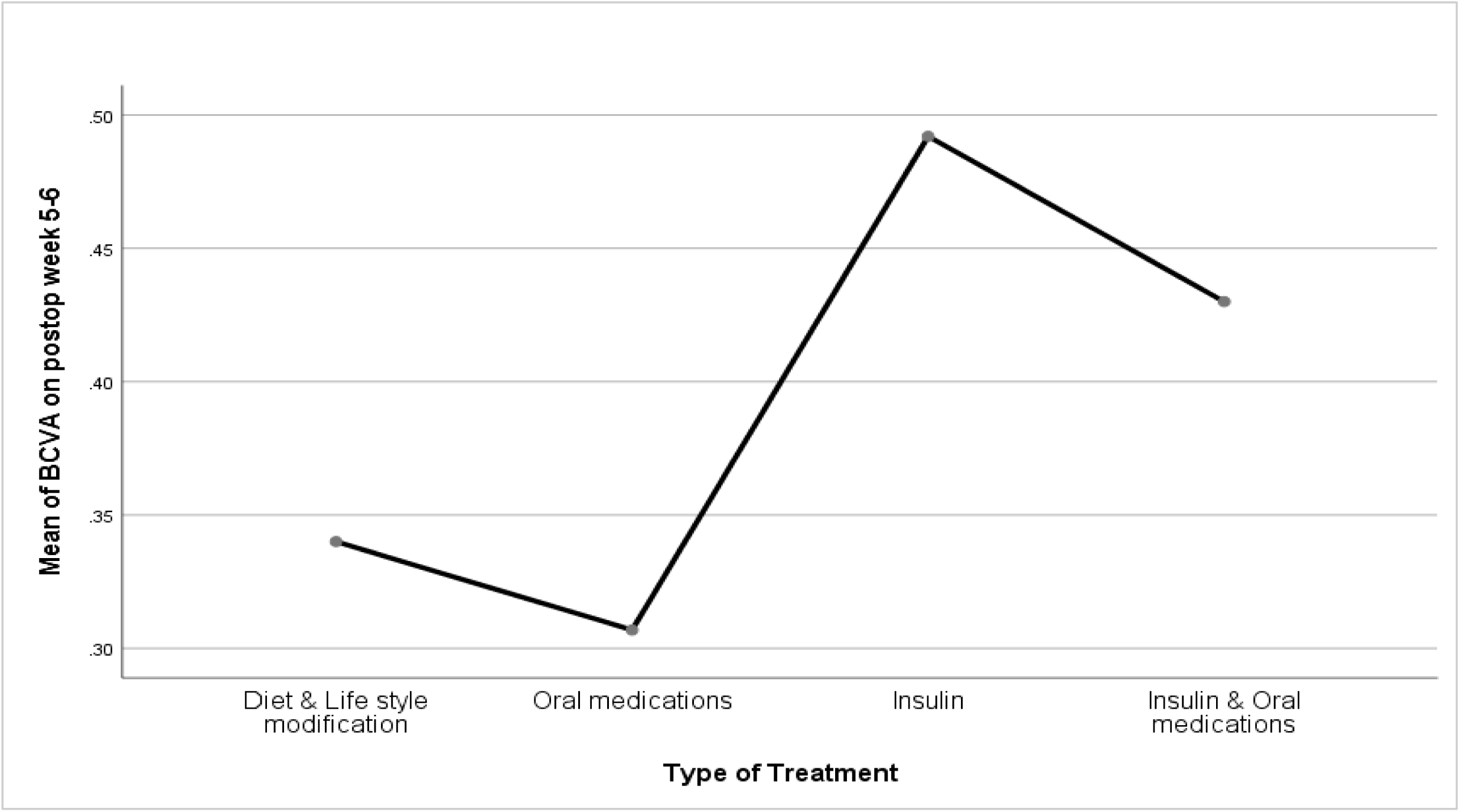
Graph showing the type of treatment for diabetes versus BCVA the 6^th^ postoperative week after MSICS in diabetic patients at the UoG Comprhensive Specialized Hospital Tertiary Eye care and training center from January to December 2024.

Out of ten eyes having borderline visual outcome, BCVA of 6/24-6/60, 9 (90%) eyes had systemic hypertension, as compared to 52.5% in those with good outcome (p=0.017).There was statistically significant difference in the final visual outcome at the 6^th^ postoperative week period between those with and with out diabetic retinopathy. Patients without DR had better mean final BCVA of 0.34±0.15 than those with mild NPDR, 0.38±0.21, (p= 0.028) and advanced PDR (1.1±0.35) with p= 0.002. Similarly, those patients with mild NPDR, moderate NPDR (0.47±0.24), and high-risk PDR (0.36) had better mean VA level than those with advanced PDR (p< 0.05).

### Intraoperative and Post operative Complications

Postoperative complications were seen in 32 (44.4%) eyes; the commonest being corneal edema/ Striate keratopathy seen in 18 (25%) eyes, most of which were mild. Posterior capsular tear (PC-tear) was seen in 6 (8.3%) eyes. (Table 6)

**Table 6.**
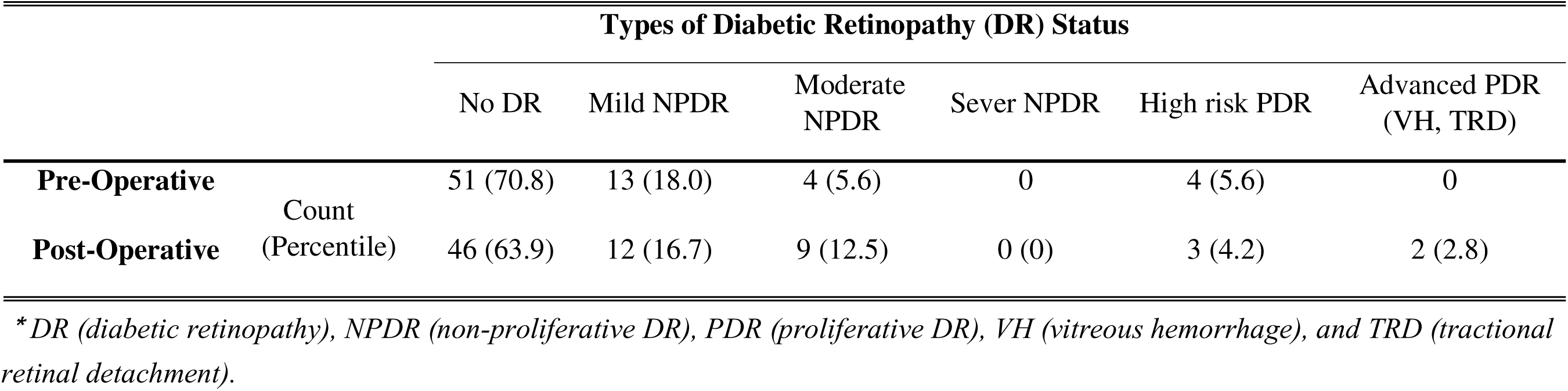
Distribution of preoperative and postoperative grades of DR in diabetes patients who undergone SICS at UoG Comprehensive Specialized Hospital Tertiary Eye Care and Training Center, January-December, 2024.

Optical Coherence Tomography (OCT) was done in 52 (72.2%) eyes on the 1^st^ postoperative week and in 46 (63.9%) eye on the 5^th^ postoperative week. From those eyes that undergone OCT exam, cystoid macular edema (CME) was found in 8 (15.4%) eyes and 3 of them were treated with intravitreal triamcinolone acetonide (IVTA), while 5 of them were followed until the 5^th^ week with topical medications and 2 of which improved, 2 remain the same, and 1 worsened in VA from 6/12 to 6/36. (Table 7)

Other postoperative complications include, retained lens material seen in 6 (8.3%) eyes, half of them underwent cortical washout but the remaining had no indication, pupillary membrane and pigment in 3 (4.2%) eyes, anterior chamber inflammation and raised intraocular pressure each in 2 (2.8%) eyes, and IOL capture/entrapment in 1 (1.4%) eye. Posterior capsular opacity (PCO) was found in 3 (4.2%) eyes and treated with Nd-YAG laser capsulotomy. (Table-7)

### Preoperative and post operative Levels of Diabetic Retinopathy (DR)

At the final follow-up examination 11 (15.3%) eyes had progressed in diabetic retinopathy, one of them had progressed two grading steps and the other one progressed three grading steps on ETDRS DR grading system (Reference). Preoperatively, 51 (70.8%) eyes had no DR, 13 (18.0%) had mild NPDR, 4 (5.6%) moderate NPDR, and another 4 (5.6%) had quiescent high-risk PDR. At the 6^th^ week follow-up period, 46 out of 51 eyes of patients without DR remained un-progressed while 4 of them developed mild NPDR, and 1 moderate NPDR. In addition, 8 out of 13 mild NPDR remained the same, 4 progressed to moderate NPDR, and 1 to advanced PDR with VH. All eyes with moderate NPDR didn’t show progression after surgery. Moreover, 3 out of 4 eyes with quiescent high-risk PDR remained the same and 1 progressed to advanced PDR (Table 8).

## DISCUSSION

This is the first prospective follow-up study in Ethiopia done on the outcomes of MSICS specifically on diabetic patients with cataract which showed MSICS resulted in improvement in vision in nearly all diabetic patients while 85% achieved post refraction good visual outcome 6 weeks after surgery.

Visual improvement in one or more lines, at the end of 6 weeks, was achieved in 70 (97.2%) eyes, remained the same in 1(1.4%) and was worse in 1(1.4%) eye. This was similar to a study done in India in which VA improved in 94.6%, remained the same in 4.5%, and was worse in 1 (0.6%) eye 26, but better than a study done in Nigeria where VA improved in 84.2%, despite similar mean BCVA of 0.38±0.21 Vs 0.38±0.49 in Nigeria. 24 The lower VA in Nigerian study may be due to insertion of average IOL as preoperative biometry was not done.

At the 6^th^ postoperative week, 54 (75%) eyes had UCVA of ≥6/18 and 61 (84.7%) had BCVA of ≥6/18, both outcomes being slightly below the WHO recommendations, and 45 (62.5%) had BCVA of ≥6/12. These results are in line with studies done in the USA 35 and India 23 where 62%, and 62.1% had VA of ≥6/12 at the end of the 4th post operative week respectively. But, lower than a study done in Austria 20 where 71% achieved VA ≥6/12, p= 0.001. In this study there was only 1(1.4%) patient who had poor visual outcome and this is better than the mentioned studies in USA 25 and Nigeria 31, where the rates of poor visual outcome were 9% and 30% respectively.

In this study, patients without DR had better post MSICS mean BCVA of 0.34±0.15 than those with DR (p< 0.05). This is in line with a study conducted in Sweden 22 where VA in eyes with DR (p=0.004) 6 weeks after surgery was worse than those with out DR. Similarly, those patients with any grade of retinopathy had better mean BCVA than those with advanced PDR (p< 0.05) 27. In a study done in USA, patients with NPDR were nearly 5 times less likely (P = .023) and patients with PDR 30 times less likely (P < .0001) to achieve good postoperative BCVA than patients without retinopathy. 10

At the final follow-up examination 11 (15.3%) eyes of our study patients had progressed in DR. This is consistent with a study done in Austria 20 in which progression was seen in 12% study eyes but no patient developed PDR in operated eye, in contrast to our study where there was 1 patient who progressed to PDR. However, studies done in Israel 21 and Korea 22 found that operated eyes had more progression of DR, 41% and 30.6% respectively, than the non-operated contralateral eyes (P <.05), which is worse than our study.

In this study, posterior capsular tear (PC-tear) was the most frequent and the only intra-operative complication seen in 6 (8.3%) eyes which was lower than a study done in Nigeria in which PC-tear was seen in 26.1% of patients. Postoperative complications, seen in 32 (44.4%) eyes, were slightly higher than a study done in Nigeria, 33% 24 but significantly lower than a study in Saudi Arabia (62.7%). 23 Corneal edema/ SK, seen in 18 (25%) eyes, was significantly better than the above studies; (62.4%) in Saudi Arabia, p< 0.05, and (30.4%) in Nigeria. Posterior capsular opacity (PCO) found in 3 (4.2%) eyes, was also better than the Nigerian study (8.7%). 24

Cystoid macular edema (CME) was found in 8 (13.4%) of our study eyes and this was similar to a study done in Sweden 25(12%) but significantly lower than studies done in Austria (31%) 20 and Israel (50%) [27], p< 0.05. Pre-existing DR was the reason for higher CME in the Israeli study. In a study done in China 6 and Sweden 27, duration, type of diabetes, level of retinopathy, and poor glycemic control were risk factors for the development of CME. However, our study didn’t show any association between CME and these factors. This might be due to the fact that none of diabetic patients in this study had poor glycemic control at the time of cataract surgery and hence this potential correlation could not be studied. In most studies, the presence of CME has been recognized as risk factor for poor (<6/60) postoperative VA. 10, 21, 24 But, in this study, 3 eyes with CME had VA of 6/36 which is in the borderline range of post MSICS visual outcome and no patient with CME had associated poor visual outcome after surgery.

Systemic Hypertension was seen in 43 (62.3%) patients of our study. A similar high incidence of hypertension, 60.9% and 71.0%, were reported in studies in Nigeria and Saudi Arabia.23,24 Over 83.3% of our diabetics in this study and with retinopathy changes had concomitant hypertension and those with hypertension tend to have lower visual outcome than those with out it (P=0.017). Similar results were seen in a study done in India 26, showing significant association between hypertension and the incidence of DR. The mean duration of diabetes mellitus prior to surgery of 10.8 years in this study was higher than 8.1 years (p=0.001) reported by Onakpoya *et al* in Nigeria*. 24*

### Conclusion and Recommendation

Although the sample size is relatively small and the duration of follow up short, this study showed that MSICS in diabetes patients with cataract resulted in an overall improvement in visual acuity and with very low rate of poor visual outcome and about 85% of operated patients archived good visual outcome after refraction. However, this finding is still slightly below the WHO recommendations, although it is still better than previous similar cataract surgical outcome studies done at the same place and other sites in Ethiopia..

## Data Availability

All data produced in the present study are available upon reasonable request to the authors

